# NeurOne: High-performance Motor Unit-Computer Interface for the Paralyzed

**DOI:** 10.1101/2023.09.25.23295902

**Authors:** Dominik I. Braun, Daniela Souza de Oliveira, Patricia Bayer, Matthias Ponfick, Thomas Mehari Kinfe, Alessandro Del Vecchio

## Abstract

We have recently demonstrated that humans with motor-and-sensory complete cervical spinal cord injury (SCI) can modulate the activity of spared motor neurons that control the movements of paralyzed muscles. These motor neurons still receive highly functional cortical inputs that proportionally control flexion and extension movements of the paralyzed hand digits. In this study, we report a series of longitudinal experiments in which subjects with motor complete SCI received motor unit feedback from NeurOne. NeurOne is a software that realizes super-fast digitalization of motor neuron spiking activity (32 frames/s) and control of these neural ensembles through a physiological motor unit twitch model that enables intuitive brain-computer interactions closely matching the voluntary force modulation of healthy hand digits. We asked the subjects (n=3, 3-4 laboratory visits) to match a target displayed on a monitor through a cursor that was controlled by the modulation of the recruitment and rate coding of the spared motor units using a motor unit twitch model. The attempted movements of the paralyzed hands involved grasping and hand digit extension/flexion. The target cursor was scaled in a way that the subjects could increase or decrease feedback by either recruiting or derecruiting motor units, or by modulating the instantaneous discharge rate. The subjects learned to control the motor unit output with high levels of accuracy across different target intensities up to the maximal achievable discharge rate. Indeed, the high-performance motor output was surprisingly stable in a similar way as healthy subjects modulated the muscle force output recorded by a dynamometer. Therefore, NeurOne enables tetraplegic individuals an intuitive control of the paralyzed muscles through a digital neuromuscular system.

**Significance Statement:** Our study demonstrates the remarkable ability of individuals with complete cervical spinal cord injuries to modulate spared motor neurons and control paralyzed muscles. Utilizing NeurOne, a software, we enabled intuitive brain-computer interactions by digitalizing motor neuron spiking activity and employing a motor unit twitch model. Through this interface, tetraplegic individuals achieved high levels of accuracy and proportional control which closely resembled motor function in intact humans. NeurOne provides a promising digital neuromuscular interface, empowering individuals to control assistive devices super-fast and intuitive. This study signifies an important advancement in enhancing motor function and improving the quality of life for those with spinal cord injuries.

## Introduction

The human hand is a remarkable structure with a complex set of movements that allow us to perform various tasks with ease. The control of hand movements is governed by a network of neural pathways that originate from the brain and the spinal cord and involve upper and lower motor neurons that control muscle forces. Electromyography (EMG) measures the electrical activity generated by muscle fibers during muscle contraction, with surface EMG (sEMG) being a non-invasive technique that can provide a comprehensive picture of motor unit activity across space and time (1, 2). Recent advancements in sEMG, particularly high-density sEMG (HD-sEMG), have allowed for accurate extraction of individual motor units using techniques such as convolutive kernel compensation (CKC) and fast independent component analysis (FastICA) (3–9). The characteristics of motor units have been investigated in both isometric and dynamic movements of the hand (4, 8, 10–13), with some studies showing the identification of unique motor units specific to certain movement patterns (14). Real-time decomposition of sEMG signals into motor unit firings, also known as online decomposition, has been successfully applied using convolutive blind source separation (BSS) techniques and gated recurrent units (GRU) (15–19).

For individuals with neuromuscular diseases or paralysis resulting in hand immobility, visual feedback of their hand movement intention is not possible. However, real-time identification of the firing motor unit activity from HD-sEMG signals might provide a solution for this lack of control. Ting et al. demonstrated that an individual with motor complete SCI still had functional motor neurons that can be extracted through the decomposition of HD-sEMG signals (20, 21). Similarly, we found unique motor units in eight motor complete SCI patients with a lesion at level C5-C6 who attempted predetermined hand movements (22). These patients were also able to track a visual cue on a monitor by modulating the discharge rate of the identified motor units in real-time (22).

Here, we present NeurOne, a software that provides paralyzed individuals with fast and accurate motor neuron feedback. As motor neurons represent the last neural code of movement that is then translated into muscle force, this interface enables direct control of the movements that were once paralyzed without the need of remapping to new motor dimensions. The software uses an online decomposition method that extracts motor unit action potentials from HD-sEMG signals through convolutive BSS embedded with a super-fast digitalization of the spiking activity (>30Hz), and a motor unit twitch model with physiological delay for the user-in-the-loop computer interaction. Although there are algorithms already capable of identifying the motor unit activity (15, 23, 24), these have very low time resolutions (<10 Hz) and do not include a realistic motor unit twitch model and therefore are not intuitive. More importantly, these previous algorithms have not been developed for paralyzed individuals which requires software with high user-in-the-loop capabilities, as demonstrated in the paragraphs below. The software is used by asking individuals with paralyzed hands to attempt various dynamic hand movements guided by a virtual hand to ensure that the HD-sEMG signals are synchronized. The HD-sEMG signals (128 electrodes) are measured from the surface of the forearm, and the extracted motor unit action potentials are used to decode the signals at a rate of 32 Hz, providing real-time feedback on task-related motor unit firings. After identifying the motor unit spike trains, NeurOne generates a task-related cumulative motor unit spike train, which is convolved with a physiological optimized motor unit twitch model to provide smooth feedback. To evaluate the accuracy of NeurOne, participants are asked to follow a requested trajectory that involves ramps with different activation levels. The accuracy is then calculated using the Pearson correlation coefficient (PCC) *r* and the root-mean-square error *RMSE* normalized to the respective activation level. We evaluated the accuracy on a subset of three patients with chronic cervical SCI who visited the laboratory over the course of up to two months. After just one day of training sessions, these patients could reliably track a visual cue on a monitor at a large range of neural activation levels. The feedback provided by NeurOne reached a coefficient of variation *cv* similar to the variability of the measured force in healthy subjects during the plateaus of ramp trajectories in different hand and lower limb muscles.

This innovative software offers a potential solution for individuals with paralysis resulting in hand immobility, providing them with a new level of control in a minimally invasive way. By allowing paralyzed individuals to use their remaining motor neurons to control their hand movements through real-time feedback, NeurOne offers a promising avenue for restoring mobility and independence.

## Results

### Interfacing Motor Units in SCI

We present a novel technique for non-invasive interfacing of the spinal motor neurons in individuals with motor and sensor-complete cervical SCI. Our method involves the application of BSS on HD-sEMG recordings to identify individual motor unit firings in real-time and rendering of the neural activity through a super-fast decomposition and integration of visuomotor feedback through a motor unit twitch model. The HD-sEMG electrodes are placed on the extensor digitorum and flexor digitorum superficialis muscle in the forearm to measure muscle activity, as these muscles are involved in almost all digit movements of the human hand.

We integrated our non-invasive motor unit interface based on convolutive BSS into our software NeurOne, which allows users to interact with physiological latency with the spared neural activity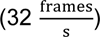. This latency enables visuomotor feedback without the perception of any delay for the user. Figure 1 shows the overview of NeurOne describing the pipeline for decoding motor unit spiking activity and the closed-loop user interaction display where SCI subjects followed predefined trajectories with a cursor controlled by their smoothed motor neuron spiking activity during hand digit movements. NeurOne decoded individual motor unit firings in real-time by decomposing the measured HD-sEMG signals on the forearm (Figure 1A).

**Figure 1.**
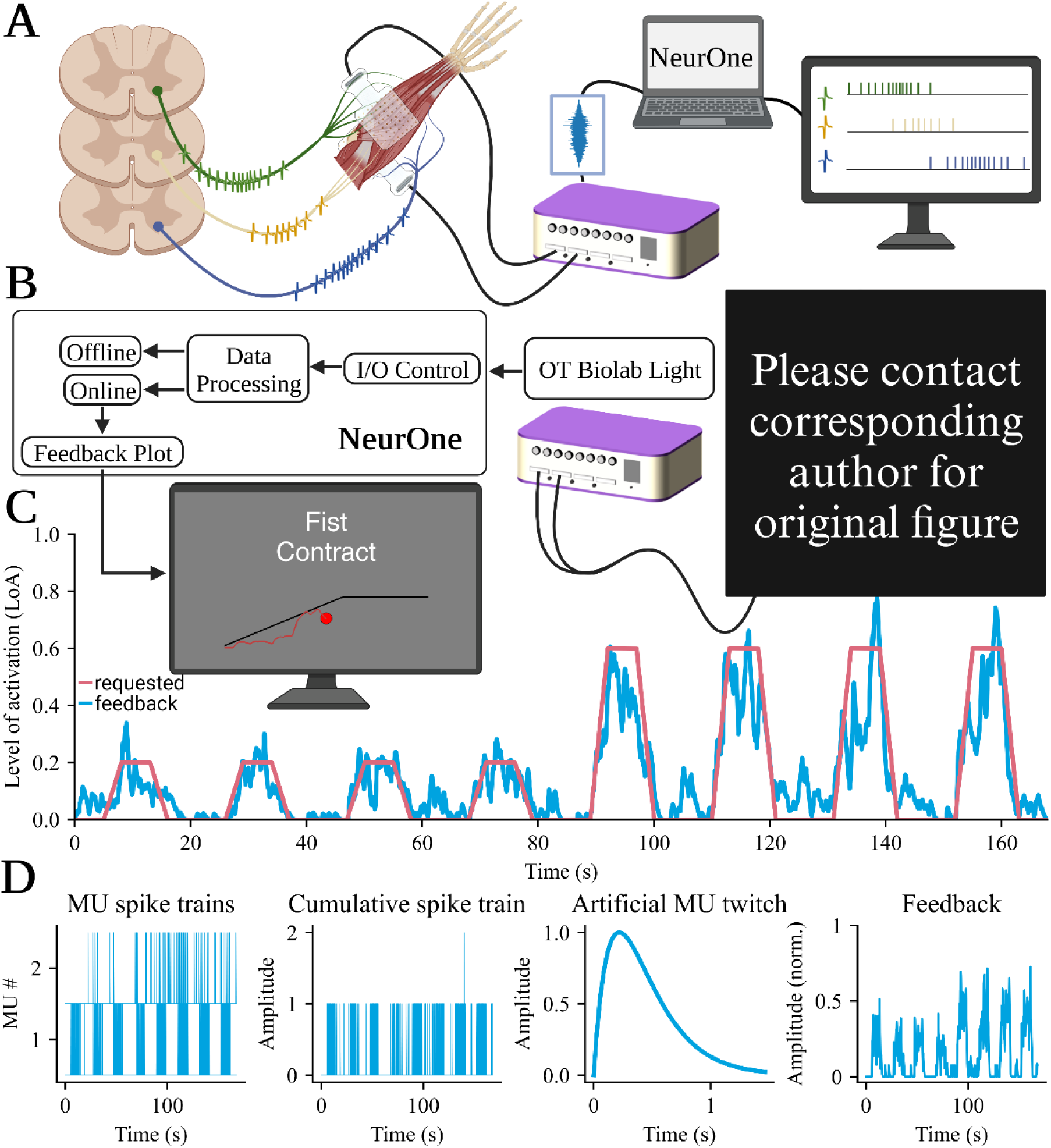
Overview of the experimental protocol used in individuals with spinal cord injury (SCI). A) We recorded high-density surface electromyographic (HD-sEMG) signals from the forearms of participants with SCI by applying two electrode grids with 64 channels each on top of the extensor digitorum and flexor digitorum superficialis muscles. These signals represent an estimate of the activity of the spared motor units that controls hand movements. We used a multichannel amplifier to collect the HD-sEMG signals and stream them to a computer that runs NeurOne. NeurOne decomposes the streamed HD-sEMG signals into individual motor unit firings. B) NeurOne used in the study where either offline or online decomposition on the acquired HD-sEMG signals from the forearm of the participant was performed. By attempting specific hand movements such as power grasp or pinch, the participants were instructed to follow a trajectory displayed on a screen during the online session. The neural feedback for the hand movements was calculated by NeurOne and displayed to the participant through a cursor on a monitor. C) An online session of participant S3, where the participant followed a requested trajectory (red line) by modulating the motor unit activity (blue line). The participants attempted to control the movement of the paralyzed hand, and the feedback from NeurOne allowed real-time adjustments of the spared motor commands to achieve the desired trajectory. D) NeurOne calculates the feedback by convolving the task-related cumulative motor unit spike train decomposed by NeurOne with a physiological optimized motor unit twitch model. This approach provides smooth and super-fast feedback that helped the participants adjusting the movements in real-time.

We performed longitudinal tests on three individuals diagnosed with complete SCI affecting motor and sensory functions in four separate sessions, over a period of two weeks, except for participant 2, who could only complete three sessions. For this participant, the first two sessions occurred within a week, while the final session took place two months later. The subjects present no movement in their hand, and they have no visible feedback when asked to attempt tasks. Here, we demonstrate that the feedback provided by NeurOne can bypass the injury and allow SCI individuals to reliably interact with a computer by attempting hand movements.

In each session, we performed a short warm-up in which the subjects were asked to follow a virtual hand displayed on a screen. Subsequently, we recorded 20 seconds of contractions to find the separation matrices (the motor unit filters), which are used in the BSS iterative process to calculate the source signals from the observations and from which the motor unit action potentials are calculated through spike-triggered-averaging. During the online part, we applied these filters such that the subjects could follow the requested trajectory with high accuracy (Fig. 2B). One online session of participant S6 is displayed in Figure 1C. The feedback is represented through the blue line while the requested trajectory is shown in red.

**Figure 2.**
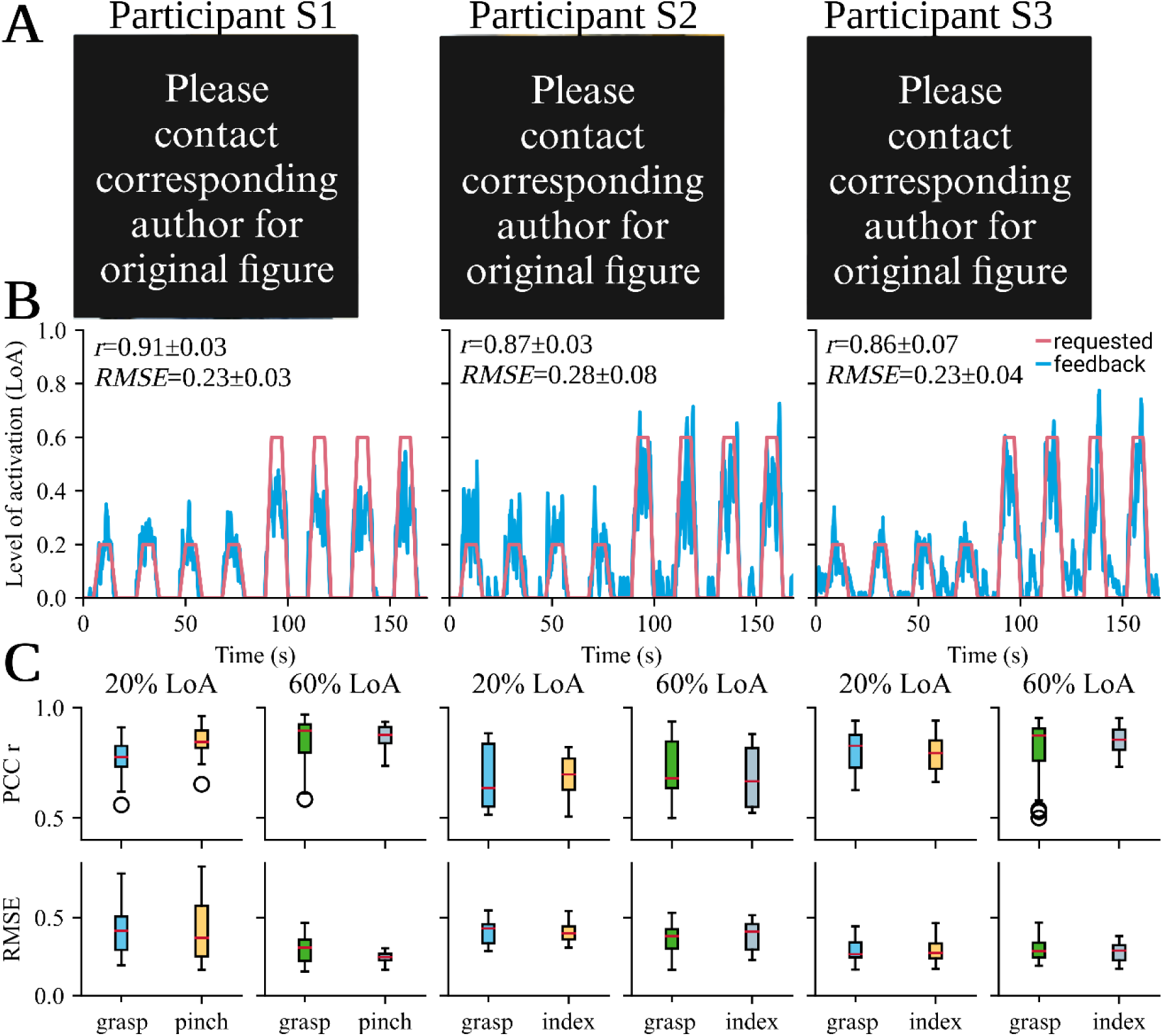
Performance of the participants in the study. A) The three participants in the study during a session. Two electrode grids, each having 64 electrodes are placed on the skin of the forearm of the paralyzed hand. After performing a warm-up and recording 20 seconds of high-density surface electromyography (HD-sEMG) the online session is performed. B) The best online attempted movements throughout all sessions (a total of nine sessions per task spanning over three training days) where the participants followed a requested trajectory (red line) consisting of eight ramps by their task-related motor unit activity (blue line). The accuracy of the performance is calculated through the Pearson correlation coefficient *r* and the root-mean-square error *RMSE* per activation. C) Correlation and error were calculated individually for each ramp/feedback pair throughout the first three training sessions for all participants shown for each task and differed between the activations of 20% and 60%. Ramp/feedback pairs that had a correlation below *r*<0.5 were discarded as they were marked as not followed. The correlation *r* and error *RMSE* demonstrated largely consistent patterns between different activation levels and tasks. However, it is noteworthy that participant S1 was the only participant showing significant differences between lower and higher activations in both metrics.

To calculate the feedback, i.e., the smoothed motor neuron spiking activity, we identified all motor unit spike trains involved in an individual hand digit movement, summed the spike trains across all motor units, and convolved the firing activity (series of zeros and ones) with an artificial motor unit twitch model (Figure 1D). The digital twitch embedded in NeurOne simulates the muscle twitch in a human muscle and has a latent period, a contraction phase, and a relaxation phase. Our approach to feedback calculation enabled high accuracy in tracking the requested trajectory (see paragraph below). We implemented the decomposition and rendering of motor unit activity by utilizing the high-performance graphical processing unit that enabled the display of the motor unit feedback and spike trains with real-time resolution 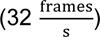. We then evaluated the performance of our feedback across the different experiments and in comparison, to intact healthy individuals, which are described below. Metrics across groups are described as mean±standard deviation.

### Accuracy of the neural feedback

All three participants with sensory and motor complete SCI were able to follow the requested trajectory with high levels of accuracy by modulating task-related motor units. The attempted tasks involved power grasp (hereafter grasp) for all participants and pinch grasp (hereafter pinch, S1)/index flexion/extension (hereafter index, S2 and S3) depending on the subject. Figure 2A shows the participants in the experimental environment with the applied HD-sEMG electrode grids. Across the first three sessions the Pearson correlation coefficient r (PCC) and the root-mean-squared error RMSE are calculated for each task and for the ramps of different levels of activations (LoA) individually for each ramp/feedback pair. The level of activation (hereafter referred simply to activation) refers to the extent of motor unit activation, i.e., motor unit discharge rate, relative to the maximal activation observed during the offline recording. Figure 2B shows the whole recording of the online session with the highest average correlation r and lowest average error RMSE per activation for each participant. The neural feedback trajectory calculated by NeurOne is displayed in blue and the requested trajectory in the red. The neural feedback trajectories of each participant follow the requested trajectory with some deviation. The average correlation r (r_1_=0.909±0.028, r_2_=0.866±0.034, r_3_=0.860±0.072; p_1, 2, r_=0.248, p_1, 3, r_=0.173, p_2, 3, r_=0.974) and error RMSE (RMSE_1_=0.231±0.031, RMSE_2_=0.280±0.081, RMSE_3_=0.228±0.042; p_1, 2, RMSE_=0.243, p_1, 3, RMSE_=0.995, p_2, 3, RMSE_ =0.208) throughout their best session for participants S1-S3 were similar, suggesting that NeurOne provides high proportionality using motor unit spiking activity.

We found a difference between the average *RMSE* of the lower (20% of maximum) and higher (60% of maximum) activations for participants S1 and S2. Specifically, for S1, we observed a significant difference (*p*=0.037) in the average *RMSE* between lower (*RMSE_1, 20_*=0.208±0.027) and higher (*RMSE_1, 60_*=0.254±0.013) activations. Similarly, for S2, a significant contrast in average *RMSE* values was evident (*RMSE_2, 20_*=0.344±0.061 vs. *RMSE_2, 60_*=0.216±0.032), with a p-value of 0.017. These results indicate that accuracy in following ramps is more difficult with lower activations than with higher activations.

Despite these RMSE variations, there were no significant differences in correlation *r* for both S1 and S2. The correlation values remained consistent for S1 (*r_1, 20_*=0.904±0.037 and *r_1, 60_*=0.914±0.011, with p-value of 0.684) and S2 (*r_2, 20_*=0.853±0.023 and *r_2, 60_*=0.879±0.037, with p-value of 0.335). In the case of participant S3, we found no significant difference between activation levels for both correlation *r* (*r_3, 20_*=0.824±0.071 and *r_3, 60_*=0.897±0.052) and error *RMSE* (*RMSE_3, 20_*=0.214±0.027 and *RMSE_3, 60_=*0.338±0.009), with p-values of 0.203 and 0.257, respectively.\

Figure 2C illustrates the overall performance across sessions. All participants displayed a robust linear relationship between task performance and activation levels, with an average correlation coefficient exceeding *r*>0.785. The correlation was significantly higher at higher activation levels (*r_20_* =0.769±0.056, *r_60_*=0.806±0.075, *p*=0.024). Notably, participant S1 exhibited a strong linear relation in both activation levels and tasks (*r_1, 20, pinch_*=0.853±0.051, *r_1, 60, pinch_*=0.867±0.051, *r_1, 60, grasp_*=0.867±0.090), except for the grasp task at 20% maximum activation (*r_1, 20, grasp_*=0.783±0.077), which was significantly lower (p-values in respect to grasp at 20% activation: *p_grasp, 60_*=1.7e-4, *p_pinch, 20_=*1.1e-3, *p_pinch, 60_*=4.7e-5). In contrast, participants S2 (*r_2, 20, grasp_*=0.696±0.158, *r_2, 20, index_*=0.684±0.106, *r_2, 60, grasp_*=0.724±0.146, *r_2, 60, index_*=0.681±0.140) and S3 (*r_3, 20, grasp_*=0.802±0.097, *r_3, 20, index_*=0.795±0.079, *r_3, 60, grasp_*=0.851±0.091, *r_3, 60, index_*=0.847±0.063) did not exhibit significant differences in their correlations between the two different activations and tasks.

Regarding error, lower activation levels had generally higher error values, while higher activation levels had lower error values (*RMSE_20_*=0.369±0.059, *RMSE_60_*=0.304±0.047, *p*=5.3e-7). Notably, participant S3 demonstrated the lowest error for lower activation levels (*RMSE_3, 20_*=0.288±0.076) and a similar error to participant S1 for higher activation levels (*RMSE_1, 60_*=0.269±0.069; *RMSE_3, 60_*=0.274±0.055). Participant S2 showed similar error values to participant S1 for lower activation levels (*RMSE_1, 20_*=0.415±0.172; *RMSE_2, 20_*=0.406±0.074). However, participant S2 exhibited the highest error for the highest activation levels (*RMSE_2, 60_*=0.365±0.094).

Participants S2 and S3 consistently maintained errors in following the requested trajectory, with no significant differences between higher and lower activations and tasks. However, participant S1, showed a significant difference between lower and higher activations and tasks (*p_grasp20, grasp60_*=0.004, *p_grasp20, pinch20_*=0.927, *p_grasp20, pinch60_*=5.42e-6, *p_grasp60, pinch20_*=0.018, *p_grasp60, pinch60_*=0.455, *p_pinch20, pinch60_*=2.92e-5). Moreover, this participant had the lowest overall error for the pinch task at 60% maximum activation (*RMSE_1, 60, pinch_*=0.246±0.034) but also the highest overall error for the grasp task at lower activation levels (*RMSE_1, 20, grasp_*=0.426±0.159) indicating that the lower activations were more difficult to follow for this participant.

Additionally, when examining the interquartile range *IQR* across all tasks and activations for correlation, participant S1 demonstrated the lowest *IQR* (*IQR_1, r_*=0.082±0.009), indicating a high level of consistency. Participant S3 followed with a slightly higher *IQR* (*IQR_3, r_*=0.112±0.031). In contrast, participant S2 exhibited a considerably larger range than the other two participants in correlation (*IQR_2, r_*=0.223±0.068). As for the calculated error RMSE between the ramp and feedback, participant S1 had the highest average range across all tasks and activations (*IQR_1, RMSE_*=0.166±0.110). However, this was mainly influenced by the higher ranges for error *RMSE* at lower activations (*IQR_1, RMSE, 20_*=0.245; *IQR_1, RMSE, 60_*=0.078) emphasising the significant differences between lower and higher activations for participant S1. On the other hand, participant S3 displayed the lowest range across all metrics, tasks, and activations (*IQR_3, RMSE_*=0.088; *IQR*_3, r_=0.112). Interestingly, for participant S3, the range for lower activations was smaller compared to higher activations (*IQR_3, RMSE, 20_*=0.101, *IQR_3, RMSE, 60_*=0.081, *IQR_3, r, 20_*=0.135, *IQR_3, r, 60_*=0.089).

These findings illuminate the consistency and variability in participants’ performance across tasks and activation levels, offering valuable insights into individual dissimilarities and patterns of response. Moreover, we observed a consistent and robust training effect for all subjects. Within just a few days of using NeurOne, the participants exhibited remarkable improvement, accurately tracking a prescribed trajectory, as described below.

### Improvement of neural feedback

Figure 3 illustrates the progress made by the participants during the three training sessions across three consecutive days that spanned over 2 weeks for participants S2 and S3. For participant 1 the first two training sessions spanned over 1 week while the last session had to be conducted two months later. Figure 3A displays the best (highest average correlation across all ramp/feedback pairs) online session for participant S2 for the index finger on each training day. On the first day, participant S2 had an average correlation *r_1_*=0.054*±*0.351 and error *RMSE_1_*=0.574±0.154 across all feedback/ramp pairs of this session. Moreover, the normalized activation levels from the neural feedback remained almost constant throughout the recording. By the second day, the neural feedback during the resting phase had become silent, and while the feedback at the requested activation of 60% did not reach 60%, the activation level for those ramps was higher than for the ramps at 20% of maximum activation. We speculate that the subjects learned to silence the motor units with tonic activity (firing when no task was displayed on the monitor) that were observed on day 1. The average correlation of *r_2_*=0.477±0.108 and error of *RMSE_2_*=0.448±0.041 was significantly improved. On the third day, participant S2 was able to modulate the feedback at the requested activation level, and the feedback trajectory tended to overshoot the requested activation

**Figure 3.**
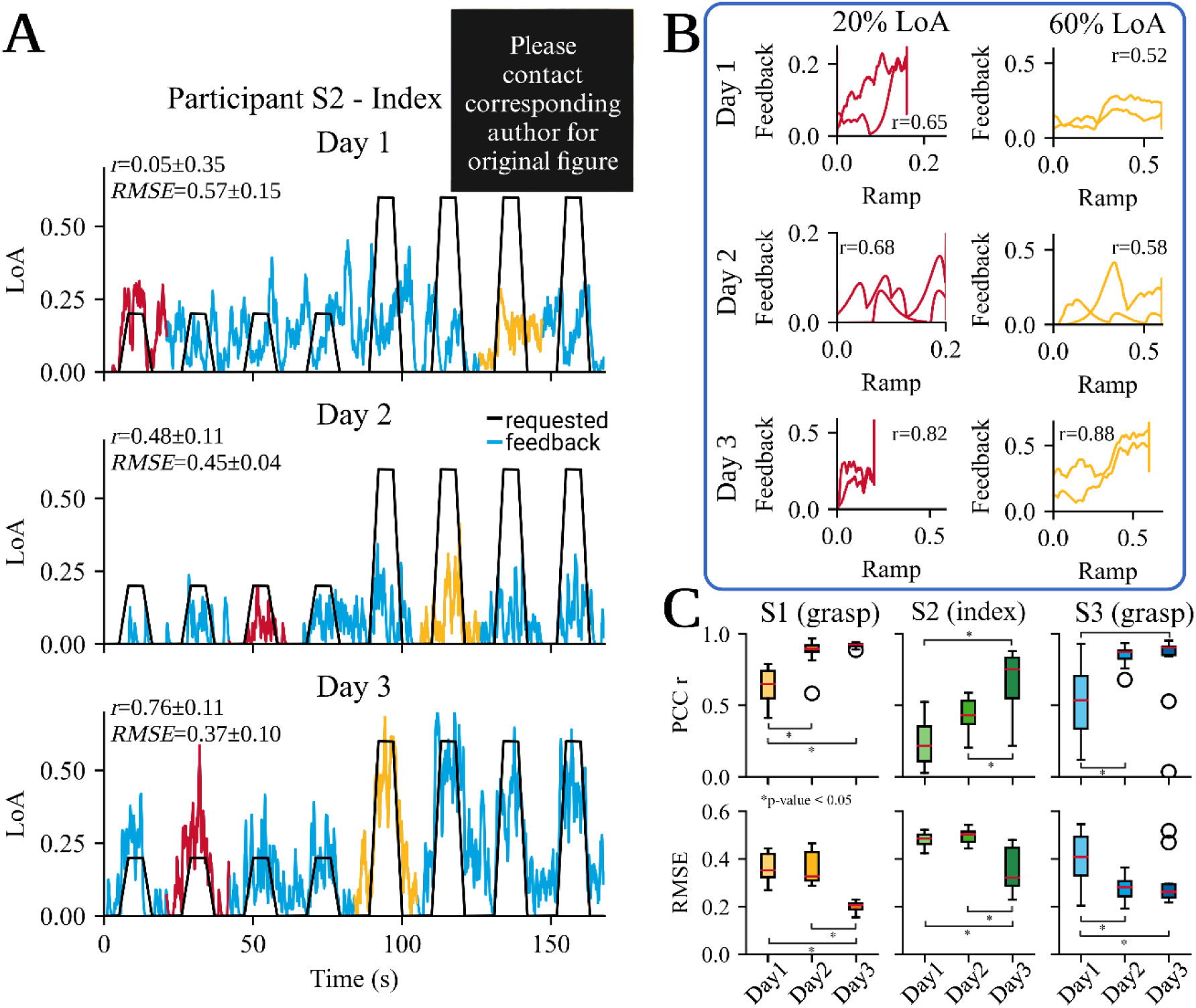
The effectiveness of the proposed neural feedback system in improving the accuracy of tracking a requested trajectory with a cursor. NeurOne was tested on three participants (S1, S2, and S3) over three training days spanning between seven days (S2) up to 2 months (S1). A) shows the improvement in proportional control of motor unit activity over time for participant S2. On the first day of training, no proportional control was observed, as feedback was activated even when not requested. However, by the second day, the participant was able to activate motor unit activity only when it was requested. On the third day, the participant was able to modulate the feedback with high proportionality and low error. B) presents the correlation and error values between the best feedback and requested trajectory for each training day for participant S2, as calculated from the best correlated feedback/ramp pair in the online recording. The plot demonstrates that the correlation improves over the course of the training days. C) Boxplots of the Pearson correlation coefficient *r* and root-mean-square error *RMSE* per activation for each participant at 60% of the maximum activation for one task. All participants showed a significant increase in the correlation *r* (Δ*r_1_=*147.6%, *p_1_*=1.33e-6; Δ*r_2_=*275.6%, *p_2_*=8.16e-4; Δ*r_3_=*172.9%, *p_3_*=2.44e-3 for participants S1, S2 and S3 respectively) and a significant decrease in the error from day 1 to day 3 (Δ*RMSE_1_=*45.6%, *p_1_*=3.54e-5; Δ*RMSE_1_=*25.6%, *p_2_*=0.011; Δ*RMSE_1_=*37.6%, *p_3_*=2.72e-3 for participants S1, S2 and S3 respectively). Participants S1 and S3 achieved consistent accuracy in following the trajectories, as the range in performance at individual ramps decreased (Δ*r_1_=*94.8%, Δ*RMSE_1_*=64.3%; Δ*r_3_=*98.6%, Δ*RMSE_3_*=66.9%) over the training sessions. In contrast, participant S2 showed an increase in the range, but the median values were higher for the correlation and lower for the error on day 3 than on the other days.

Although the neural feedback was still active during the resting phase, the activation was much lower than during the actual ramps. During the best performance of the index finger task, the participant had an average correlation of *r_3_*=0.759±0.109 and an error of *RMSE_3_*=0.372±0.104 across all feedback/ramp pairs of this session. Across the days, participant S2 was able to improve the proportional control of the cursor by more than 1,400% and reduced the error by 35.2%. Figure 3B shows the requested activation level plotted against the feedback calculated by NeurOne and displays the differences between the days more clearly. For each day, a ramp/feedback pair with the highest correlation value was selected for both 20% and 60% activations. The feedback at 20% of maximum activity is colored red, while the feedback at the requested activation level of 60% is colored yellow. The activations at 20% on day 1 showed a negative correlation of *r_1, 20_*=-0.20 for 20% and *r_1, 60_*=-0.14 for 60% of the maximum activation. By day 2, the correlation for the target activation level of 20% reached *r_2, 20_*=0.68 and for 60% the correlation had the value *r_2, 60_*=0.34. By day 3, the correlation for both activation levels reached *r_3, 20_*=0.78 for 20% and *r_3, 60_*=0.88 for 60% of the maximum activation level.

Across three days of training, all participants demonstrated a higher correlation and lower error in at least one task when the activation level was set to 60%. Figure 3C illustrates the performance of the ramp/feedback pairs, which revealed that on the first day, each participant had a lower correlation (*r_1, day1_*=0.626±0.141; *r_2, day1_*=0.246±0.215; *r_3, day1_*=0.525±0.284) and higher error (*RMSE_1, day1_*=0.362±0.063; *RMSE_2, day1_*=0.479±0.042; *RMSE_3, day1_*=0.402±0.105). On the third day, all participants showed a significant increase in correlation values (*r_1, day3_*=0.924±0.016, *p*=1.33e-6; *r_2, day3_*=0.678±0.208, *p*=8.16e-4; *r_3, day3_*=0.908±0.003, *p*=2.44e-3) and a decrease in error values (*RMSE_1, day3_*=0.197±0.028, *p*=3.54e-5; *RMSE_2, day3_*=0.357±0.094, *p*=0.011; *RMSE_3, day3_*=0.251±0.032, *p*=2.72e-3). Compared to participant S2, participants S1 and S3 achieved high correlation values by the second day (*r_1, day2_*=0.899±0.042, *p_1_*=9.67e-7; *r_3, day2_*=0.861±0.047, *p_3_*=7.82e-4). However, the error was not reduced for participant S1 (*RMSE_1, day2_*=0.353±0.066, *p*=0.938). Overall, there was an increase of 147.6%, 275.6%, and 172.9% in the correlation and a decrease of 45.6%, 25.6%, and 37.6% in the error for participants S1, S2, and S3, respectively.

Furthermore, the interquartile range *IQR* in the results decreased for participants S1 and S3 from day 1 to day 3. For participant S1, the range in correlation decreased by 94.8% (*IQR_1, day1, r_*=0.192 to *IQR_1, day3, r_=*0.010) and in error by 64.3% (*IQR_1, day1, RMSE_*=0.098 to *IQR_1, day3, RMSE_*=0.035). Although the range decreased significantly for correlation after one day of training, the error was only reduced on the third day. For participant S3, the range for correlation and error decreased after the first day (*IQR_3, day1, r_*=0.369 to *IQR_3, day2, r_*=0.053; *IQR_3, day1, RMSE_*=0.163 to *IQR_3, day2, RMSE_*=0.054). From day 1 to day 3, the interquartile range *IQR* in correlation decreased by 98.6% (*IQR_3, day3, r_*=0.005) and in error by 66.9% (*IQR_3, day3, RMSE_=*0.054). However, only participant S2 showed an increase in range and correlation values, but the error values decreased. This was particularly evident in the error range, which was similar on the first two days (*IQR_2, day1, RMSE_*=0.040 to *IQR_2, day2, RMSE_*=0.044), but increased by 400% on the last training day (*IQR_2, day3, RMSE_*=0.160). Regarding the correlation, there was a decrease of 32.8% in the range between the first two days (*IQR_2, day1, r_*=0.244 to *IQR_2, day2, r_*=0.164), but on the last training day, the correlation range was significantly increased (*IQR_2, day3, r_*=0.284).

### Validation of NeurOne

Figure 4 depicts the software architecture of NeurOne, including the feedback calculation process for achieving seamless and ultra-fast feedback delivery to the user. The interface to the amplification device, which records the HD-sEMG signals, enables the streaming of 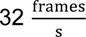 with a sampling frequency of 2048 Hz (64 data samples per frame) for a total of 408 channels. Offline decomposition of a 20-second recorded HD-sEMG signal (49,960 data samples per channel) was completed in 3:05±0:10 minutes. During online decomposition, the measured time difference between two frames was *t_Δframe_*=31.3±0.42 ms, resulting in an average of 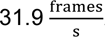. The measured time to calculate the feedback was *t_calc_*=3.07±0.7 ms. Updating the plot windows for the spike trains and feedback took *t_plot_*=4.33±0.7 ms after the frames were received. Participants in the study did not report any delay between the attempted task and the displayed feedback.

**Figure 4.**
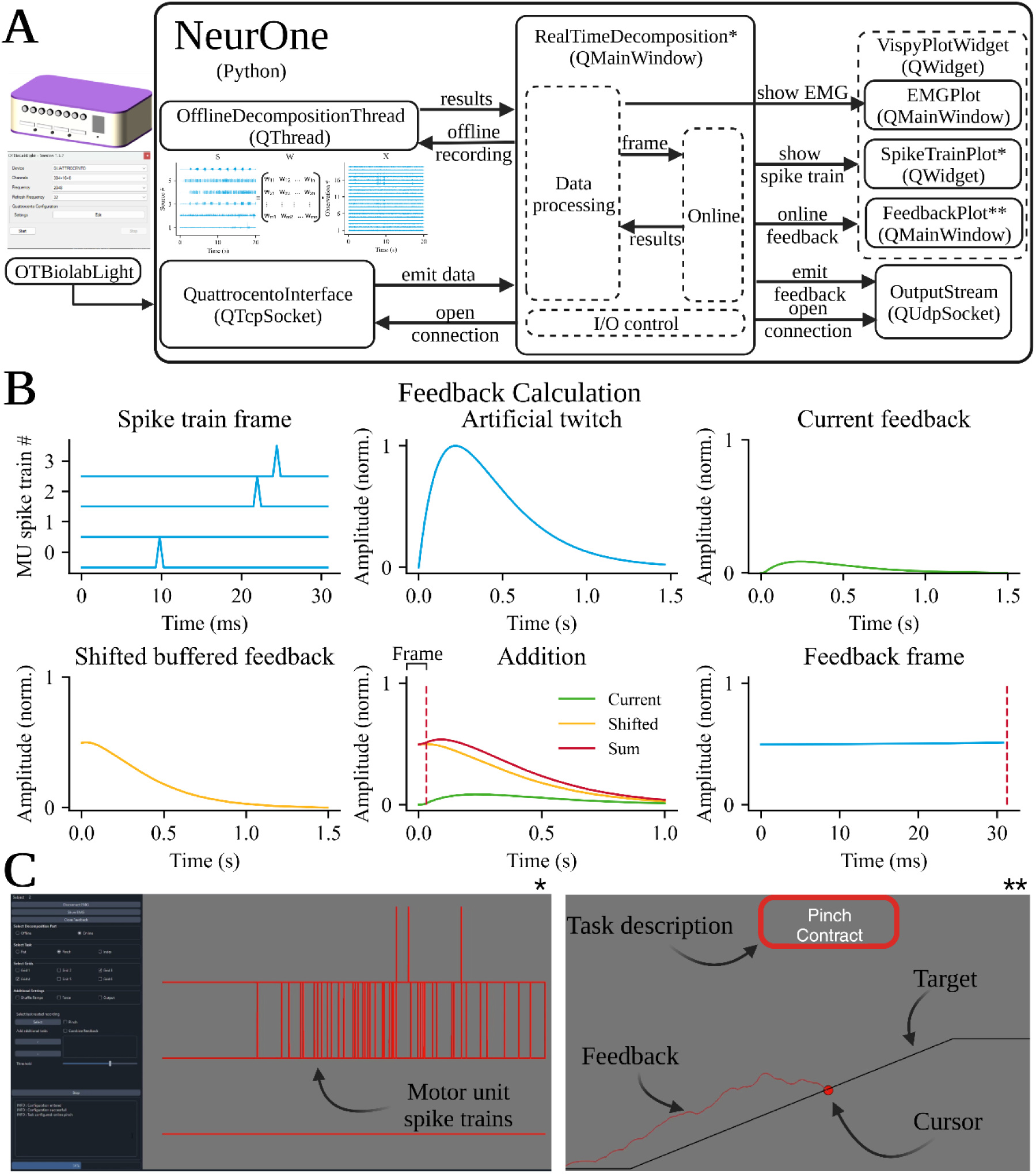
Overview of NeurOne’s software architecture and the feedback calculation process displayed to the participants. A) The software is utilizing the PySide6 Python module and uses a QuattrocentoInterface (based on QTcpSocket) to communicate with the amplification device software *OT Biolab Light*. This data is then sent to the main window of NeurOne, which handles the graphical user interface (GUI), motor unit spike train plots, and data processing. NeurOne can perform either offline or online decomposition of incoming data. The spike trains of all motor units, including those of the main and sub-tasks, are displayed in the main window using the SpikeTrainPlot widget, while the calculated feedback is plotted in a separate FeedbackPlot window (based on QMainWindow), making it possible to display the monitor specifically for the participant in a dual monitor setup. NeurOne can also display the high-density surface electromyographic signals in real-time using the EMGPlot window (based on QMainWindow). NeurOne also provides the functionality of streaming the calculated feedback through an object of the OutputStream class (based on QUdpSocket), which maps the feedback of the selected task on the involved fingers to control a virtual hand or mechatronic systems. B) The feedback calculation that enables fast and smooth feedback for controlling the cursor to track the requested trajectory. The identified spike trains of the task-related motor units are summed up into a cumulative spike train, which is then convolved with a motor unit twitch model. The induced feedback from this frame is then added to the calculated feedback from previous frames. From the resulting summed feedback, the first 64 samples, i.e., 31.25 ms (red-dotted line), are taken as the feedback frame. The average of the feedback frame is mapped on the cursor. C) Main window of NeurOne’s GUI that displays the identified motor unit spike trains in real-time (left) and the feedback window that is displayed to the participants of the study (right). NeurOne’s main window allows users to choose tasks, electrode configurations, online and offline parts. In the case of the online part, users can select one main task from which the feedback is displayed in the feedback window and additional sub-tasks. The real-time decoded motor unit spike trains are displayed in the main window, with tasks being colored differently. The feedback window, displayed to the participants in the study, provides task instructions and displays the cursor (red dot) representing the current feedback frame and its history (red line) while the user is asked to follow the requested trajectory (black line) by attempting the pinch task.

Figure 5 illustrates the validation process of the feedback calculation algorithm integrated in NeurOne. As previously described, this algorithm uses a motor unit twitch model to smooth the discharge rates of motor unit firings. To validate this approach, the convolutive feedback method was applied to decomposed motor unit spike trains of 22 healthy individuals acquired by previously conducted experiments (10 subjects in the first (10, 25) and 12 subjects in the second experiment (26, 27)) in an offline analysis. In the second experiment of the study (first experiment with healthy individuals), the force exerted by the index finger during an isometric contraction was measured using a mechanical apparatus, while HD-sEMG signals were recorded from the third dorsal interosseous (FDI) muscle using a 64 channel electrode grid (Figure 5A/B). The second experiment (second experiment with healthy subjects) involved the measurement of HD-sEMG and force during isometric ankle dorsiflexion. Two HD-sEMG electrode grids with 64 channels each were placed on the skin above the musculus tibialis anterior (TA). The force was measured using an ankle dynamometer.

**Figure 5.**
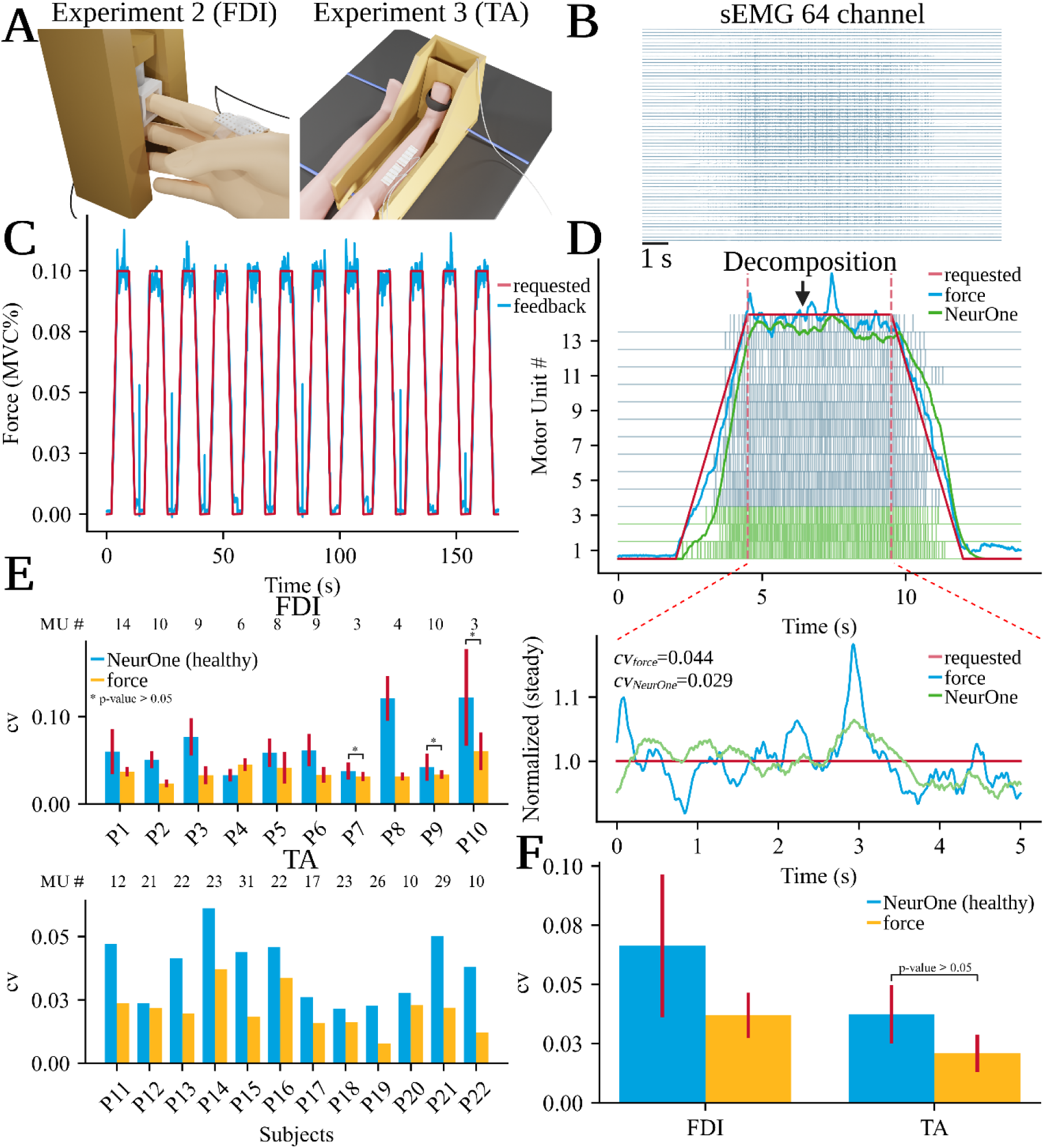
Procedure used to validate the feedback calculation method of NeurOne. A) Two experiments were conducted that involved placing high-density surface electromyography (HD-sEMG) electrode grids consisting of 64 channels on the first dorsal interosseous (FDI) muscle (left) and the musculus tibialis anterior (TA, rechts) of 23 healthy subjects (10 and 12 in experiment 2 and 3 respectively). At the same time, the isometric force produced during index finger abduction (FDI) and ankle dorsiflexion (TA) was measured through a mechanical apparatus. B) A recorded HD-sEMG signal during a ramp contraction of experiment 2 (14 seconds) was analyzed and decomposed into motor unit spike trains. C) The subjects were instructed to follow a specific trajectory with their generated force, consisting of twelve ramps with a target activation level of 10% of maximum voluntary contraction (MVC). The requested trajectory is displayed with the red line and the force feedback measured with the blue line (displayed for experiment 2). D) The cumulative spike train of the three motor units (green) from the recorded HD-sEMG signal during a ramp contraction of the index finger abduction task were used in the feedback calculation approach of NeurOne. Additionally, the requested trajectory (red), the force signal (blue), and the feedback calculated by NeurOne (green) are displayed. Four seconds of the plateau part of the ramp (between the vertical dotted red lines) were extracted for each signal and experiment and normalized on its mean. Furthermore, the coefficient of variation *cv* was calculated for the presented ramp plateau. E) The mean and standard deviation of the coefficient of variation *cv* were calculated for each participant of experiment 2 (FDI, P1-10) and 3 (TA, P11-22) across all ramps. The coefficient of variation *cv* was displayed for the output of NeurOne’s feedback calculation method (blue bars) and the recorded force signals (yellow bars) for the healthy subjects. F) Average coefficient of correlation *cv* across all participants for experiment 2 and 3.

In both experiments, the healthy individuals were instructed to follow a predetermined force trajectory. For the experiment with the FDI, the requested force trajectory was represented by twelve ramps with a target activation level of 10% of their maximum voluntary contraction (MVC), presented in Figure 5C. The MVC was determined prior to the study and one isometric ramp contraction in the requested trajectory had a duration of 14 seconds. The second experiment with the ankle dorsiflexion had a target force of 35% MVC with incline and decline of 5%/s and involved only one ramp.

The HD-sEMG signals recorded in both experiments were decomposed into motor unit spike trains, and the three most active motor units were selected for the validation process. NeurOne’s feedback was derived from analyzing the cumulative spike train of a specific subset of identified motor units, which was then compared with the recorded force signal (Figure 5D). To simulate the number of identified motor units in real-time experiments involving SCI, we carefully selected a subpool of motor units consisting of the three motor units with the highest number of firings during the contraction phase. The coefficient of variation *cv* of NeurOne’s feedback was evaluated to determine its similarity to the coefficient of variation *cv* of the measured force signal. Therefore, the steady parts of the reference signal and NeurOne’s feedback were extracted and normalized on the mean of their respective steady part. The coefficient of variation *cv* of the force signal in one ramp of the first experiment (FDI) was found to be *cv_force_*=0.044, while the coefficient of variation *cv* of NeurOne’s feedback was *cv_NeurOne_*=0.078.

Figure 5E presents an overview of the average coefficient of variation values obtained from the protocol ramps of experiment 2 (FDI) and 3 (TA). While the coefficient of variation value for the reference signal in experiment 2 (FDI) was generally lower than the coefficient of variation from NeurOne’s feedback calculation, three healthy participants showed almost similar coefficients of variation values (cv_P7, force_=0.031±0.005, cv_P7, NeurOne_=0.038±0.010, p=0.079; cv_P9, force_=0.034±0.005, cv_P9, NeurOne_=0.042±0.016, p=0.105; cv_P10, force_=0.060±0.022, cv_P10, NeurOne_=0.122±0.056, p=0.072). Participant P4 exhibited an even lower coefficient of variation value with NeurOne’s feedback than with the recorded reference signal (cv_P4, force_=0.045±0.007, cv_P4, NeurOne_=0.033±0.007, p=6.58e-4). In experiment 3 (TA) all subjects had a slightly higher coefficient of variation for the calculated motor unit feedback (NeurOne). Three subjects (P12, P18 and P20), however, showed an almost similar coefficient of variation *cv* to force (*cv_P12, force_*=0.022, *cv_P12, NeurOne_*=0.024; *cv_P18, force_*=0.016, *cv_P18, NeurOne_*=0.022; *cv_P20, force_*=0.023, *cv_P20, NeurOne_*=0.028). However, some subjects (P11, P13-15, P19, P21-22) had a much higher coefficient of variation *cv* for the motor unit feedback of NeurOne compared to the measured force (*cv_P11, force_*=0.024, *cv_P11, NeurOne_*=0.047; *cv_P13, force_*=0.020, *cv_P13, NeurOne_*=0.041; *cv_P14, force_*=0.037, *cv_P14, NeurOne_*=0.061, *cv_P15, force_*=0.018, *cv_P15, NeurOne_*=0.044; *cv_P19, force_*=0.008, *cv_P19, NeurOne_*=0.023; *cv_P21, force_*=0.022, *cv_P21, NeurOne_*=0.050; *cv_P22, force_*=0.012, *cv_P22, NeurOne_*=0.038). Across all healthy subjects during the index finger abduction task, the coefficient of variation value was *cv_FDI, NeurOne_*=0.066±0.030 for the feedback calculation method implemented in NeurOne. In comparison, the coefficient of variation *cv* for force (*cv_FDI, Force_*=0.037±0.005) was significantly lower (Δ*cv*=44%, *p*=3.51e-3) and exhibited greater consistency with a narrower range across subjects. This contrasted with the coefficient of variation *cv* observed during ankle dorsiflexion, which was generally lower than during the index finger abduction task. Noteworthy, when utilizing NeurOne, the coefficient of variation *cv* (*cv_TA, NeurOne_*=0.038±0.012) achieved a similar value with no significant differences compared to the force during experiment 2 (*p*=0.10) and experiment 3 (*cv_TA, Force_*=0.021±0.008, *p*=0.120). These findings suggest the effectiveness of NeurOne in providing comparable results to force measurements in both experiments.

These small discrepancies between the measured force and the rendered force by NeurOne are related to numerous factors which include a small number of motor units that were used for the analysis, offline experiments, and other nonlinear characteristics of motor neuron to muscle force generation. However, the differences in actual force and digitally rendered force by NeurOne were very small and negligible (Figure 5 D-F).

## Discussion

In this study, we introduce NeurOne, a non-invasive and intuitive software that provides users with immediate neural feedback on the spared motor unit activity, which enabled three SCI individuals to train and control the spared neural activity after many years of motor complete paralysis. We presented the framework behind NeurOne which consists of two main parts. We then evaluated NeurOne on longitudinal experiments and proved that this framework enables SCI individuals to control a cursor on a screen in a similar way as intact healthy individuals modulate the isometric force output.

The first part of the framework is the offline decomposition that tries to find suitable filters that extract the source signals, i.e., the motor unit firings convolved with their motor unit action potentials. The offline decomposition method, which was adapted for NeurOne, a convolutive blind source separation algorithm, is extensively tested and validated against iEMG by different researchers (4, 15, 19). The decomposition method is performed fully automatically and requires only 3 minutes and 5 seconds (3:05±0:10) to complete. This makes it considerably faster than comparable solutions (15).

The online decomposition with the intuitive motor unit interface for the paralyzed is the second and novel part of NeurOne. It applies the found filters from the offline decomposition, i.e., the separation matrix *W,* the motor unit action potentials and the maximum value of the calculated feedback of the cumulative offline spike train on the streamed HD-sEMG frame. After identifying the motor unit firings, the task-related cumulative spike train is used to calculate a smooth and super-fast feedback by convolving it with a motor unit twitch model.

Our study demonstrated that NeurOne provides highly effective feedback, enabling participants with paralyzed hands to accurately follow a requested trajectory with strong proportionality (correlations of *r*=0.91/0.87/0.86) and minimal error (*RMSE*=0.23/0.28/0.23 for participants S1/2/3) across an entire online recording consisting of eight ramps during attempted hand movements. Note that during these movements the subjects show no movements of the hands (see Ting et al. and Oliveira et al. for more details on this finding (21, 22)). Furthermore, our results revealed that NeurOne was capable of motivating and engaging participants to track the requested trajectory more accurately over the course of multiple training days. For example, participant S2 showed a substantial improvement in proportionality (*r*=0.05 to *r*=0.76) and a reduction in error (*RMSE*=0.57 to *RMSE*=0.37) for the index task over three training days. It should be noted that the reported correlation and error values are averaged across all eight consecutive ramps in an online recording, and therefore do not imply that participants were unable to follow any ramp in the first online sessions. Variability in correlation and error exhibited greater variation during the initial training sessions. This suggests that as participants became more familiar with the system, their ability to consistently and accurately track trajectories improved. This training phenomenon highlights the promising utility of NeurOne, which has a direct connection to spinal motor neurons, in the field of neural rehabilitation for people with paralysis.

Consistency in control signals is crucial for the effective use of NeurOne, particularly in applications involving mechatronic systems such as exoskeletons or prostheses. Individuals with neuromuscular conditions or paralyzed limbs need a control system that feels natural, and NeurOne can provide smooth and fast feedback that can be modulated proportionally to different activation levels within the same time window. The participants achieved an almost similar (no significant differences for participants S2 and S3, participant S1 has a significant difference for the grasp task at the lower activation) correlation for both activations across tasks (above *r*>0.79 in average) indicating a strong proportionality between the voluntary motor unit spiking activity and the requested trajectory. Especially in applications where high durability is crucial, a strong proportionality between voluntary motor unit spiking activity and target level, along with low error, becomes vital. This is because maintaining a constantly high activation level would lead to exhaustion and muscle soreness.

However, there are also limitations to the proposed feedback calculation, particularly regarding the normalization of feedback. The MVC is typically used for this purpose but cannot be calculated using force sensors in patients with SCI who are not able to produce force with their hands. To address this, we engaged participants as much as possible during the offline phase through dynamic contractions and used the maximum value of the calculated offline feedback as the MVC for normalization. However, there are differences between the online and offline spike detection methods used in our study, which we plan to address in future studies by using consistent detection methods.

The speed of the feedback calculation and presentation emphasizes the importance of timely feedback for individuals with SCI, as they do not have visible feedback of their muscle contractions. Moving average filters are often employed to smooth the discharge rate of motor units for offline and real-time presentation (15, 22). However, using such filters involves buffering the data, leading to delays in feedback presentation. In related works, this delay goes up to 500 ms due to the need to wait for four frames of data at a streaming frequency of 8 Hz (15). Additionally, the low streaming frequency results in a delayed feedback presentation, with the plot being updated only eight times per second.

NeurOne addresses these limitations by offering a high streaming frequency of 32 Hz, which is significantly higher than any previous real-time decomposition approaches (15–17, 19), and introduces significant latencies to the user. The proposed feedback calculation method using a motor unit twitch model does not require waiting for a specific amount of time, thereby eliminating the delay in feedback presentation (15).

To validate NeurOne’s feedback method based on the digital motor unit twitch model, a comparison in the variability of the signal, i.e., the coefficient of variation *cv* during the plateau phase of isometric ramp contractions in healthy subjects was conducted. In general, the coefficient of variation *cv* of the force signals was lower than for smoothed motor unit spiking activity. One reason for the higher variability observed in the participants is that force feedback was used as a reference to track the ramp trajectory on a screen, which allowed participants to gauge the steadiness of the force signal. Moreover, the number of motor units used was limited to the three most active motor units imitating the number of motor units that were found in individuals with SCI. Together with a high variability in the number of motor units identified per subject in the decomposition process, this is a limiting factor in a fair comparison with force measurement as in force generation are up to hundreds of motor units involved. Another factor that may contribute to higher variability in NeurOne’s feedback is the challenge of reliably identifying small motor units that are generally better in precise and smooth movements compared to bigger motor units. However, the small motor units are often suppressed by bigger motor units because of their bigger motor unit action potentials making it difficult for current decomposition methods to decompose the small motor units (4, 5, 8). Despite these differences, the variability of NeurOne and the measured torque level was negligible, which confirms the high robustness of the method for digitalizing motor units in SCI.

Nevertheless, a few subjects displayed a similar coefficient of variation *cv*, with one subject (P4) showing even lower variability. This finding is remarkable, given that the human muscle twitch is optimized for smooth control, resulting in low variability in measured force. This suggests that NeurOne’s feedback is also able to provide real-time smoothness and can be applied to control assistive devices.

An alternative and frequently employed method to control assistive devices, as opposed to the motor unit twitch model, is the integration of a musculoskeletal model. However, it’s important to note a difference in the torque output bandwidth between musculoskeletal models and actuators of mechatronic systems. Actuators exhibit a broader torque bandwidth when compared to musculoskeletal models. Therefore, through the normalization of NeurOne’s output, we can efficiently utilize the complete motor bandwidth, leading to improved performance.

## Conclusion

In this study, we demonstrated the efficacy of NeurOne, a noninvasive and intuitive software that provides immediate neural feedback on the spared motor unit activity of individuals with SCI. Developed with the specific goal of improving the lives of individuals who have paralyzed hands, NeurOne can help them gain greater control over assistive devices and facilitate communication. By providing real-time, high-speed, and smooth neural feedback, NeurOne enables individuals with long-term complete motor paralysis to gain real-time control of their motor unit activity and accurately track a requested trajectory with a cursor. Our findings suggest that the accuracy of tracking can be improved through training, indicating the potential for NeurOne to enhance the rehabilitation process. In addition, we performed offline analysis to validate NeurOne’s feedback by applying it to motor unit spike trains that were decomposed with a high level of accuracy during isometric index finger abduction and ankle dorsiflexion tasks in healthy participants. We observed that NeurOne’s feedback achieved a level of variability during the plateau phase of the ramps that was partially similar to the generated force. The smoothness and accuracy of the smoothed motor unit discharge rate through NeurOne support the possibility of using this software for assistive device control such as exoskeletons. Overall, our results highlight the promising potential of NeurOne to revolutionize the way individuals with paralysis interact with the world around them and improve their quality of life.

## Materials and Methods

This study involved the recruitment of three participants diagnosed with chronic motor complete SCI for experiment 1 (SCI subjects). The study employed the following criteria to select participants: (1) injury level ranging from C4-C6, (2) age between 18 and 60 years old, and (3) absence of voluntary movement of one hand or both hands. Participant S3 exhibited voluntary hand movement in their left hand. An overview of the paralyzed participants is shown in Table 1.

**Table 1.** Characteristics of recruited participants in the study.

| Subject | Age range (years) | Gender | Injury level | AIS | Sensory level* | Wrist movement | Time since injury (years) |
| --- | --- | --- | --- | --- | --- | --- | --- |
| S1 | 31-35 | M | C5 | B | C5 | yes | 9.1 |
| S2 | 36-40 | F | C5 | A | C5 | yes | 24.2 |
| S3 | 56-60 | M | C5 | A | T3 | no | 6.9 |

\* The sensory level corresponds to lowest level with normal sensory function.

**Table 1.** Characteristics of recruited participants in the study
| Subject | Age<br>range<br>(years) | Gender | Injury<br>level | AIS | Sensory<br>level* | Wrist<br>movement | Time<br>since<br>injury<br>(years) |
| --- | --- | --- | --- | --- | --- | --- | --- |
| S1 | 31-35 | M | C5 | B | C5 | yes | 9.1 |
| S2 | 36-40 | F | C5 | A | C5 | yes | 24.2 |
| S3 | 56-60 | M | C5 | A | T3 | no | 6.9 |

22 healthy subjects were recruited for experiment 2 (*index finger abduction*, 10 subjects) and experiment 3 (*ankle dorsiflexion*, 12 subjects). All procedures were conducted in accordance with the Declaration of Helsinki and were approved by the Ethical Committees of Friedrich-Alexander-Universität (approval no. 22-138-Bm, experiment 1), Imperial College London (approval no. 18IC4685, experiment 2) and University Rome ‘Foro Italico’ (approval no. 44 680, experiment 3). Prior to participation, all subjects provided written informed consent. Some data from this study have been previously published (10, 25–27).

### Experiment 1 (spinal cord injury)

The first experiment comprised multiple sessions for each participant, with S2 and S3 undergoing training on four separate days and S1 on three days. The last session for S1 occurred two months after the previous sessions, which were conducted within a two-week timeframe. During the sessions, we trained participants to enhance their neural control over two distinct tasks, utilizing an online decomposition approach to analyze a HD-sEMG signal obtained from the placement of 128 HD-sEMG electrodes on the forearm’s skin. The training tasks consisted of a power grasp, which involved the flexion and extension of the whole hand, and a pinch grasp that required the involvement of the thumb and index finger (for S1) or single-digit movement of the index finger (for S2 and S3). Participant 3 only attempted the training with the power grasp task in the first session.

Prior to commencing the training, participants underwent a one-minute warm-up in which they followed a virtual hand attempting the task in a relatively slow (0.5 Hz), sinusoidal pattern displayed on a monitor. Subsequently, we recorded a 20-second signal during which participants were asked to attempt the requested task dynamically, i.e., flexion and extension in repetition. The task was attempted dynamically during the recording because the decomposition had a higher accuracy at finding motor units compared to isometric tasks. The recorded signal was then decomposed offline to determine the unique motor unit action potentials and separation matrix *W* for the online phase. If decomposition was not finding filters or the filters were insufficient, decomposition results of the same tasks in previous sessions were selected.

Once offline decomposition was completed, we initiated the online phase, which comprised three sets, with each set including eight trajectories, and a one-minute break between sets. The trajectories consisted of ramps with increasing (three seconds) and decreasing (three seconds) flanks, as well as a plateau (five seconds). A ten-second resting phase separated each ramp. The first four ramps had an activation level of 20%, while the subsequent four ramps had an activation level of 60%. This difference in activation levels was intended to determine whether participants with paralyzed hands can voluntarily modulate their motor unit activity to match two significantly different target levels. Moreover, by having the ramps reach two different activation levels, we were able to test the proportionality at different modulating rates. The relatively long sloping parts with a duration of three seconds ensured a large period of proportional tracking. The total duration of one set was 2:48 minutes.

In each session for each task, the protocol included a warm-up period followed by the offline decomposition phase and the online training segment. Between the completion of one task and the commencement of another, a larger break of three minutes was provided. Altogether, the training per day took approximately 40-45 minutes.

### Experiment 2 (index finger abduction)

The full details of this experiment have been described previously (10, 25). We also provided a brief explanation of the methods here. A chair, table, and computer monitor constituted the experimental setup, where participants (nine men and one woman) assumed a comfortable seated position. Their dominant hand was supported by a custom apparatus, with the forearm immobilized and positioned between pronation and supination. The index finger and thumb were aligned along the forearm’s longitudinal axis, and a monitor situated 60 cm away displayed the applied force. Force measurements of the index finger and thumb were captured using a three-axis force transducer (Nano25, ATI Industrial Automation), which underwent digitization at 2048 Hz (USB-6225, National Instruments) and underwent low-pass filtering at a cutoff frequency of 15 Hz. HD-sEMG signals were obtained from the first dorsal interosseous (FDI) and thenar muscles (flexor pollicis brevis and abductor pollicis brevis) using flexible electrode grids featuring 13x5 electrodes with a 4 mm interelectrode spacing and amplified with a multichannel amplifier (Quattrocento, OT Bioelettronica; 16-bit A/D converter, bandwidth 10-500 Hz). Next, the HD-sEMG signals were processed using a well-established BSS algorithm to decompose them into individual motor unit spike trains (5, 6).

Participants engaged in force-matching tasks, involving simultaneous abduction of the index finger and flexion of the thumb, for a duration of 60 seconds. Visual feedback was provided via a moving dot cursor on the monitor, with the x-axis representing thumb force and the y-axis representing index finger force. Participants were instructed to maintain the force signal within 10% of the target for each applied force.

Prior to the tasks, MVC recordings were performed, and two 60-second trials were conducted with 30 seconds of rest between them. The experimental design aimed to explore the extent of common synaptic inputs among sets of motor neurons, requiring participants to exert forces in the same sagittal plane for both muscle sets, necessitating approximately 10 minutes of practice.

### Experiment 3 (ankle dorsiflexion)

The full details for this experiment have been described previously (26, 27). We also provided a brief explanation of the methods here. The experimental setup consisted of a custom-made ankle ergometer (OT Bioelettronica, Turin, Italy) fixed to an examination table using adjustable straps. Twelve recreationally active young men participated in the study, with their dominant leg secured to the ergometer using straps (approximately 3 cm width) at the foot, ankle, and knee. Force signals were recorded using a force transducer (CCt Transducer s.a., Turin, Italy), amplified (200 x), and sampled at 2048 Hz using an external A/D converter (Quattrocento, OT Bioelettronica, Turin, Italy). Visual feedback was provided via a custom LabVIEW software (LabVIEW 8.0; National Instruments, Austin, TX, USA) displayed on a monitor positioned 1 m away from the participants. HD-sEMG signals were recorded from the TA muscle using two semi-disposable adhesive grids, each with 64 electrodes (13x5 electrodes with an IED of 8 mm, OT Bioelettronica). The signals were sampled at 2048 Hz, bandpass filtered (10-500 Hz), and digitally converted using a 16-bit A/D converter. The HD-sEMG signals were then similar to experiment 2 processed using a well-established BSS algorithm to decompose them into individual motor unit spike trains (5, 6).

Participants underwent a standardized warm-up, consisting of eight isometric contractions of the dorsiflexors at varying intensities (4 × 50%, 3 × 70%, 1 × 90%), separated by 15–30 seconds. After the warm-up, they performed three or four MVCs with 30 seconds of rest in between. The highest MVC force determined the maximal voluntary force (MVF) used to set target forces (35%, 50%, and 70% of MVF) for subsequent submaximal contractions. Participants later performed trapezoidal contractions, gradually increasing to the target force, maintaining it for 10 seconds, and then linearly decreasing back to the resting force at the same rate. Two trials were conducted for each target force in randomized order and 3–5-minute rest intervals.

### Evaluation of experiment 1 (spinal cord injury)

The analysis of the training sessions was carried out using Python 3.11, where each ramp/feedback trajectory pair of the online recordings was partitioned and evaluated individually. The trajectories were then categorized into 20% and 60% activation levels for further analysis. To evaluate the accuracy of each ramp/feedback trajectory pair, two metrics were used, namely Pearson correlation *r* and root-mean-square error *RMSE*. Pearson correlation measures the correlation between the requested (ramp) and actual (feedback) trajectories, indicating the degree of proportionality between the two. Additionally, the error provides a measure of accuracy by assessing the distance between the requested and actual trajectories.

To enable comparison between participants, the initial three sessions were selected from participants S2 and S3, resulting in 36 ramp/feedback pairs for each task and activation level. To demonstrate the overall performance of participants during the online sessions (Figure 2C), ramp/feedback pairs with a correlation value below 50% were discarded. Boxplots were then used to plot the ramp/feedback pairs for each task, activation level, and participant, with the box representing the IQR and the median displayed as a red line. The whiskers extending from the box represent the minimum and maximum values of the data that fall within 1.5 times the IQR from the first and third quartile, respectively. The range of the data was described by reporting the IQR, as well as the mean and standard deviation of the ramp/feedback pairs. The mean and standard deviation were also calculated across all ramp/feedback pairs within the online recording, which included eight ramp/feedback pairs.

To evaluate training improvement, only ramp/feedback pairs with a positive correlation were considered. Similar to the general performance, the Pearson correlation coefficient *r* and the root-mean-squared error *RMSE* were used to describe the accuracy of the tracking. For each training day, twelve ramp/feedback pairs were evaluated. The best online recordings of participant S2 were selected based on the highest correlation values across the entire online recording. Mean and standard deviation were reported for the entire online recording, and the ramp/feedback pairs selected for the correlation plots (Figure 3B) were based on the highest correlation values within the online recording.

### Evaluation of experiment 2 (index finger abduction) and 3 (ankle dorsiflexion)

The decomposed motor unit pulses and the measured reference signal (i.e., the force) of the healthy participants were used to validate the feedback calculation approach proposed in our study. To do this, we calculated the coefficient of variation *cv*, which is the ratio of the standard deviation 𝜎(𝑥) to the mean of the reference signal *x* during the steady plateau of the trajectory:

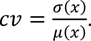

Additionally, we calculated the feedback offline, instead of online as in experiment 1, by convolving the decomposed spike trains of the three most firing motor units with the motor unit twitch model. Three motor units were selected as this is the average number of motor units identified in our study in people with SCI. Afterwards, we extracted the steady part and calculated the coefficient of variation *cv*. For each participant, we calculated the mean and standard deviation for feedback and reference signal (force) across the ramps (twelve ramps per subject for experiment 2 and the best ramp at 35 % MVC for the subjects in experiment 3). In experiment 2 the ramps that didn’t show three individual motor units spiking during the plateau phase were discarded.

### High-density surface electromyography recording

During all sessions of experiment 1, we placed two HD-sEMG electrode grids, each containing 64 electrodes, on the shaved and cleaned skin of the forearm. The electrode grids utilized in our investigation were square in shape, with an 8x8 configuration of electrodes, and an interelectrode distance (IED) of 10mm (GR10MM0808, OT Bioelettronica, Turin, Italy). To ensure consistent electrode placement, we positioned one electrode grid above the extensor digitorum and the second above the flexor digitorum superficialis, both aligned with the ulna bone. To further enhance reproducibility, we recorded the exact electrode positions by capturing images. To affix the electrode grids to the skin, we used bi-adhesive foam layered between the grids and the skin, filled with conductive paste (SpesMedica, Battipaglia, Italy), and secured them to the forearm using tape.

The HD-sEMG signals were recorded using a multichannel amplifier with 16-bit A/D conversion (Quattrocento, OT Bioelettronica). We used the OT Biolab Light software (OT Bioelelettronica) to record the signals in monopolar mode, with a sampling frequency of 2048 Hz, and filtered by a bandpass of 10-500 Hz. 408 channels were streamed in real-time using a Transmission Control Protocol/Internet Protocol (TCP/IP) with a streaming frequency of 32 Hz. However, only the 128 channels holding the HD-sEMG signals were extracted and used from the streamed data.

### Online decomposition

The first part of the online decomposition process aiming at the decoding of HD-sEMG signals into individual firings of motor units in real-time, involved an offline decomposition. The offline decomposition is necessary to determine the filters that will be applied during the second part of the process in real-time. Therefore, we conducted a recording of a dynamic task (grasp or pinch/index finger flexion/extension) and decomposed the recorded HD-sEMG signals.

The approach of the decomposition (offline and online) is based on the theoretical model of measured HD-sEMG signals. The HD-sEMG signal is a convolutive mixture of motor unit spike trains and action potentials. In matrix form it is described as:

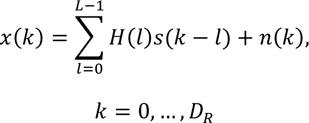

where *x(k) = [x_1_(k), x_2_(k), …, x_m_(k)]^T^* is the vector comprising all recorded observations (HD-sEMG channels) *m* and *s(k) = [s_1_(k), s_2_(k), …, s_n_(k)]^T^* is the vector comprising the spike trains of all motor units *n*. Matrix *H(l)* has the size *m* x *n* for each sample *l* and carries the information of the motor unit action potentials. *L* is the duration of the action potentials. Furthermore, *H(l)* is assumed to be constant during the recording of observations. The additive noise vector *n(k) = [n_1_(k), n_2_(k), …, n_m_(k)]^T^* comprises the noise for each observation. *D_R_* is the duration of the recording of the observations. By applying BSS techniques to this mixed model, the sources, i.e., the individual motor units, can be decomposed. For those algorithms, we assume that the identified sources are not fully correlated and are either sparse or independent (4). The algorithm that we were using for the offline decomposition of the HD-sEMG was based on the proposed convolutive BSS approach of Negro et al. (4). To reduce the noise in the observations we applied a Butterworth bandpass filter (20-500 Hz) to remove noisy frequencies where the observations are not significantly represented and a 50 Hz notch filter to remove power line interference. Following the filtering, we performed convolutive sphering as described by Negro et al. (4). The convolutive sphering method involves extension, centering, and whitening of the HD-sEMG signal. We used an extension factor of *R*=10 as we were looking for *n*=32 sources by using *m*=128 channels and an estimated action potential length of *L*=40 samples by following the general equation for the extension factor *R* (4):

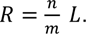

The convolutive sphering is followed by applying FastICA, which is a fixed-point iteration algorithm that maximizes the number of uniquely identified sources, i.e., the mixture of the motor unit spike trains convolved with its action potential, by using Gram-Schmidt Orthogonalization. Through FastICA, a separation matrix *W* is obtained and by multiplying it with the extended HD-sEMG signal 𝑥^*(k)* it results in the source signals *s(k)*:

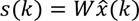

A silhouette score-based K-means driven approach is used to detect spikes from identified sources. The sources are squared, resulting in innervation pulse trains (IPTs). The peaks of the IPTs are divided into two classes: high peaks and small peaks. The small peaks, representing noise, are discarded. High peaks with a silhouette score of 0.9, indicating the distance between spike and noise, are considered as firing a motor unit.

Apart from the optimized separation matrix *W* obtained through FastICA, other results from the offline decomposition are also used in the online part. By calculating the spike triggered average (STA) for each source, we can find the motor unit action potentials to use in the real-time decomposition as templates for template matching.

To normalize the feedback calculated during the online part, a reference value is required. Without normalization, an estimation of the activation is not possible. Therefore, feedback using the offline spike trains is calculated. The feedback is the convolution of the cumulative spike train of all motor units that are found with an artificial motor unit twitch, which is simulating a muscle twitch during neural input-based contraction in humans. The real-time detection of spikes from individual motor units constitutes the second part of the online decomposition, and the pipeline is described in Figure 6. The observations x(k) in this phase consists of the streamed HD-sEMG frame (128 channels x 64 samples), which is extended and centered similar to the offline decomposition but not whitened due to high computational costs. The extended observations 𝑥^(k) are then multiplied with the separation matrix W determined during the respective task in the offline decomposition to obtain the identified sources in real-time (Figure 6A). In order to detect spikes in the current frame, the sources are subject to thresholding with a threshold value T set at 10 times the noise level, which is calculated in the first frame during rest by taking the average of each source signal in this frame (Figure 6B). However, thresholding alone may not be sufficient for reliable spike detection due to the lack of filtering of noise through a Butterworth bandpass filter in real-time compared to offline decomposition. To enhance the algorithm’s reliability, we used template matching (Figure 6C). In template matching, spike triggered action potentials of each source were correlated to the motor unit action potential extracted during offline decomposition. If the correlation between the template and the signal exceeds 0.6, the spike was accepted as valid. Subsequently, the spikes in the current frame are convolved with the artificial motor unit twitch used in the offline part. However, since the twitch length is significantly longer than the actual frame, the leftover signal is buffered for the next frames to prevent an unstable feedback signal. The feedback from the previous frames is then shifted and added to the convolutive result in each iteration.

**Figure 6.**
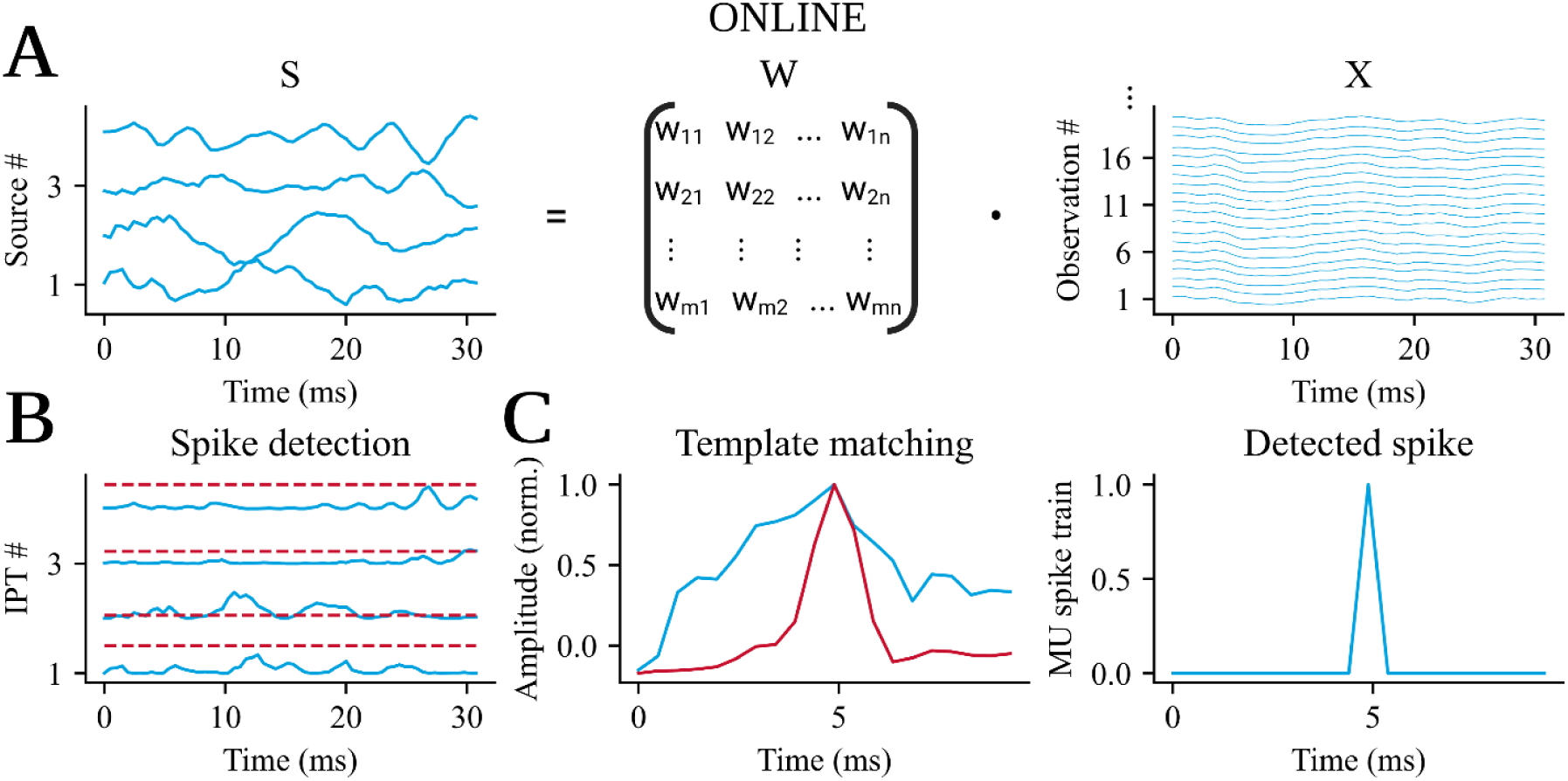
This figure depicts the online decomposition method that is utilized in our software, NeurOne. The process involves three steps. A) First, the source signals are identified in real-time by applying the separation matrix W, which was discovered in the offline stage on the extended and centered high-density surface electromyographic (HD-sEMG) signals of the current frame. B) Next, a spike detection technique is applied to the identified sources. This method detects the peak in the innervation pulse trains (IPTs), which are the squared source signals of this frame. If the peak of the IPT is greater than 10 times the noise level, it is designated as a possible spike. C) Finally, template matching is conducted to verify whether the possible spike is a motor unit firing or not. To achieve this, a window is implemented around this possible spike in the source signal and then correlated with the motor unit action potential that was identified in the offline stage. If the correlation coefficient r_threshold_ > 0.60, the spike is identified as a motor unit firing.

### Graphical user interface

NeurOne is a software that provides a GUI for real-time display of identified motor unit firings and neural feedback. Figure 4A shows the architecture of the back-end of NeurOne. NeurOne is written in Python 3.10 and utilizes the PySide6 module which provides access to the complete Qt 6.0+ framework. The RealTimeDecomposition class, which is a child class of QMainWindow, integrates the GUI and the back-end, and manages the flow of data within NeurOne for processing and plotting.

The study, in which NeurOne was used, involved recording and amplifying HD-sEMG signals from the participant’s forearm using a multichannel amplification system (Quattrocento, OT Bioelettronica, Italy). The communication between the recording software (OT Biolab Light) and NeurOne was established via TCP/IP network communication using the QuattrocentoInterface class. Depending on the selected part (offline or online decomposition), the input frame is either directly decomposed in real-time or buffered for offline analysis after recording. The online processed HD-sEMG frame, which is the motor unit spike train, is displayed in the SpikeTrainPlot widget. Furthermore, the feedback is calculated by the convolution of the motor unit spike trains with the motor unit twitch model (Figure 4B) and displayed in the FeedbackPlot window. These visualizations are based on the VispyPlotWidget class, which uses the graphical processing unit (GPU) to render the data. This is enabled by the VisPy library in Python. Furthermore, the EMGPlot is a separate window that can be opened and configured to display the streamed HD-sEMG signals and is based on the VispyPlotWidget too. Additionally, NeurOne uses the OutputStream class to open a UDP socket to stream the calculated feedback as a control signal to control a virtual hand or assistive devices. After the offline and online recordings, the results are stored in a NumPy file (.npy extension) along with a timestamp and subject identifier for subsequent data processing.

NeurOne’s GUI is shown in Figure 4C. The user can connect to the HD-sEMG measurement system, display streamed data in real-time, and start the neural interface to follow requested trajectories with the feedback cursor. Furthermore, the selection of the offline or the online part is enabled through radio buttons. The user can configure the HD-sEMG by selecting respective channel ports that are connected to the electrodes placed on the forearm and repeat the offline part until the identified filters are reliable and provide great accuracy in the online part. Tasks available to the user are grasp and pinch as well as index flexion/extension. In the online part, the user can choose filters for the main task by choosing the respective folder in the operating system’s filesystem. The main task determines which task the subject should attempt to follow the requested trajectories. Additionally, sub-tasks may be selected, whose motor unit firings are displayed alongside the main task, but without real-time display of the feedback. The requested trajectory that is used for the online protocol has four ramps with a low activation of 20% followed by four high ramps with an activation of 60%. The requested trajectory and the corresponding feedback trajectory were displayed on the FeedbackPlot window, which was located on a second monitor in front of the participants.

The evaluation of the computing and plotting time was done on a mobile laptop (XMG NEO 15 E21, Ryzen 9 5900HX, NVIDIA RTX 3080 mobile, 32 GB Ram), on which 15 motor units were recorded and visualized during the measurement. The display of spike trains and feedback had a window of 5 seconds and 128 channels of HD-sEMG were decomposed.

### Statistical Analysis

In this study, we conducted statistical analyses to investigate significant differences in the measured results using one-way ANOVA type 2 (for more than two groups) with the anova_lm function from the Python package Statsmodel and t-test (for two groups) with the ttest_ind function from the Python package Scipy.

We employed the significance level α=0.05 to determine whether there are significant differences between groups. P-values below the significance level indicate the rejection of the null hypothesis,highlighting observable significant differences. Conversely, p-values above the significance level indicate no difference in the data. To identify specific group differences after the one-way ANOVA, we conducted a pairwise Tukey test using the pairwise_tukeyhsd function from the Python package Statsmodel. In Experiment 1, we applied the statistical analysis to detect differences between lower and higher activations and between different tasks. The correlation coefficient *r* and error *RMSE* were used individually as dependent variables to assess their significance. Additionally, the analysis was used to highlight significant improvements over the training days. In Experiment 2, the statistical analysis aimed to identify significant differences between the coefficient of correlation *cv* (dependent variable) of the feedback calculated using the method implemented in NeurOne and the recorded force. Moreover, we conducted a statistical analysis across all participants in Experiments 2 and 3 to investigate significant differences in the variability of force and motor unit feedback.

## Acknowledgments

This study was partly funded by d.hip (Digital Health Innovation Platform), a cooperation between Siemens Healthineers, Medical Valley, University Hospital Erlangen, and Friedrich-Alexander University Erlangen-Nuremberg, and the German Aerospace Center (DLR) as part of the MYOREHAB project.

## Data Availability

All data produced in the present study are available upon reasonable request to the authors. An executable of NeurOne can be found <u>here</u>^1^.

## Footnotes

1 Link: https://github.com/NsquaredLab/NeurOne

## Notes

### Competing Interest Statement

The authors have declared no competing interest.

### Clinical Trial

This study is not a clinical trial.

### Author Declarations

Ethical Committee of Friedrich-Alexander-Universitaet Erlangen-Nuernberg gave ethical approval for experiment 1 in this work (approval no. 22-138-Bm). Ethical Committee of Imperial College London gave ethical approval for experiment 2 in this work (approval no. 18IC4685). Ethical Committee of University Rome 'Foro Italico' gave ethical approval for experiment 3 in this work (approval no. 44 680).

